# Screening rare genetic diagnoses for amenability to bespoke antisense oligonucleotide therapy development: a retrospective cohort study

**DOI:** 10.1101/2025.02.10.25321921

**Authors:** David Cheerie, Marlen C. Lauffer, Logan Newton, Kimberly Amburgey, Danique Beijer, Bushra Haque, Brian T. Kalish, Margaret Meserve, Rachel Y. Oh, Amy Y. Pan, Miriam Reuter, Michael J. Szego, Anna Szuto, N=1 Collaborative, Annemieke Aartsma-Rus, Michelle M. Axford, Ashish R. Deshwar, James J. Dowling, Christian R. Marshall, Zhenya Ivakine, Matthis Synofzik, Timothy W. Yu, Gregory Costain

## Abstract

**Purpose:** To estimate the proportion of molecular genetic diagnoses in a real-world, phenotypically heterogeneous patient cohort that are amenable to antisense oligonucleotide (ASO) treatment.

**Methods:** We retrospectively applied the N=1 Collaborative’s VARIANT (**V**ariant **A**ssessments towa**r**ds El**i**gibility for **An**tisense Oligonucleotide **T**reatment) guidelines to all diagnostic variants found by clinical genome-wide sequencing at a single pediatric hospital in 532 patients over a 6-year period. Variants were classified as either “eligible”, “likely eligible”, “unlikely eligible”, or “not eligible” in relation to the different ASO approaches, or “unable to assess”.

**Results:** In total, 25 unique variants across 26 patients (4.9% of 532 patients) were eligible or likely eligible for ASO treatment at a molecular genetic level, via canonical exon skipping (4), splice correction (3), or mRNA knockdown (18). Only eight of these molecular genetic diagnoses were made within a year of symptom onset. After considering disease and delivery related factors, 11 diagnoses were still considered candidates for bespoke ASO development.

**Conclusion:** A meaningful proportion of genetic diagnoses identified by genome-wide sequencing may be amenable to ASO treatment. These results underscore the importance of timely diagnosis, and the proactive identification and accelerated functional testing of genetic variants amenable to ASO treatments.

## INTRODUCTION

Rare genetic conditions are major contributors to pediatric morbidity and mortality, but few have specialized treatments.^1,2^ Increased access to genome-wide sequencing (GWS) is replacing diagnostic odysseys with therapeutic odysseys.^1,3^ Antisense oligonucleotides (ASOs) are short, synthetic chains of nucleotides that have the potential to target any gene product of interest.^4^ ASOs are versatile in their usage, as they can be employed to: (i) modulate splicing (induce exon skipping or exon inclusion, or correct aberrant (cryptic) splicing), (ii) downregulate transcripts in the case of toxic gain-of-function (GoF) and dominant negative variants, and (iii) increase protein expression from the wildtype allele.^4,5^ ASOs can be delivered most efficiently to the central nervous system (including the retina) via local injection, and the liver after systemic treatment. These are tissues commonly impacted by genetic conditions.^4–6^ At present, 16 ASOs have received U.S. Food and Drug Administration approval to treat genetic conditions (Supplemental Table 1). Proof-of-concept also exists for individualized genetic interventions with ASOs that are customized for specific genetic variants.^7–10^ However, the proportion of molecular genetics diagnoses that might be amenable to treatment with a customized ASO therapy in a real-world phenotypically heterogeneous clinical cohort is unknown.

**Table 1.**
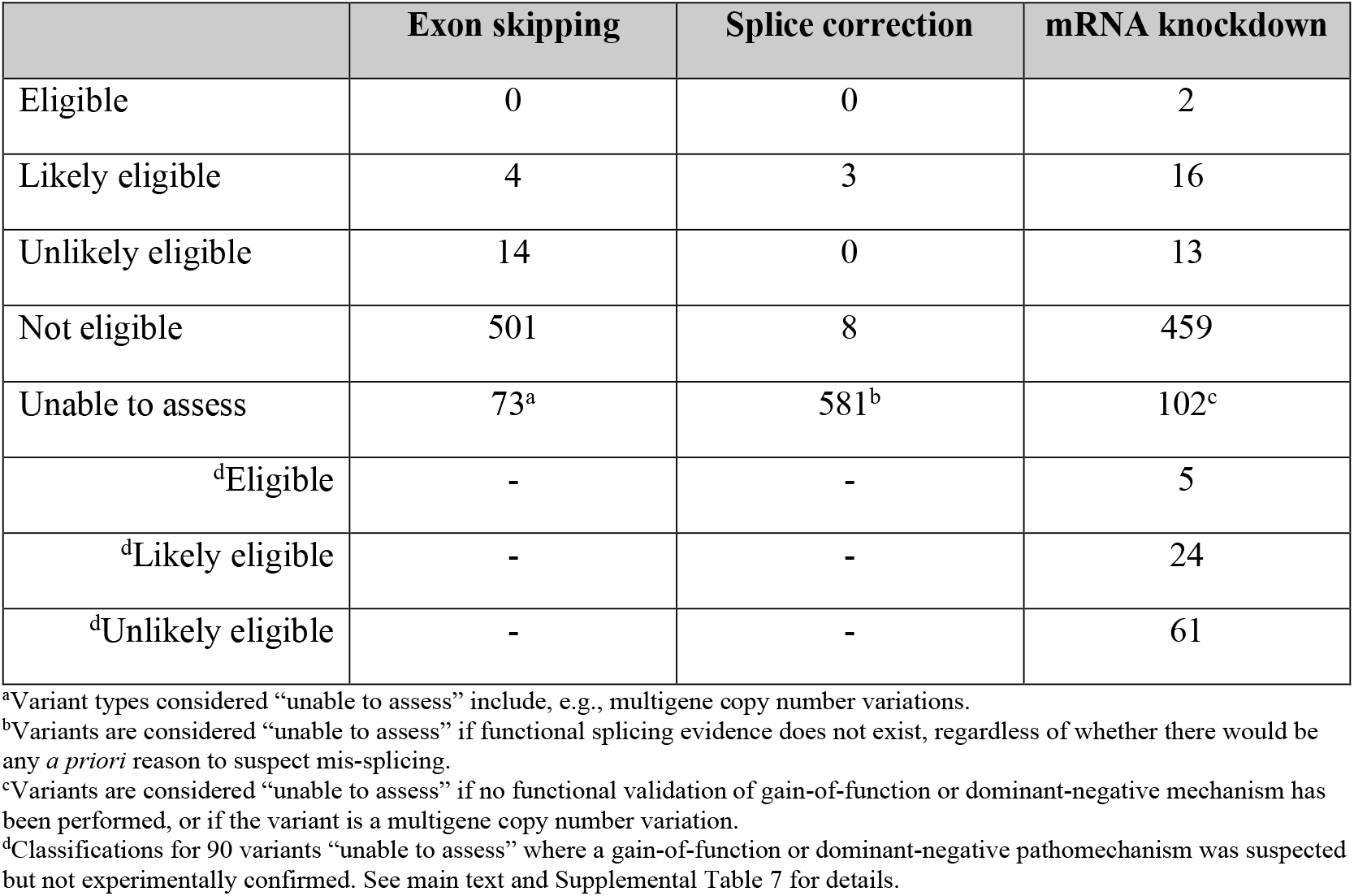
Overview of results from applying the N1C VARIANT guidelines to a set of n=592 diagnostic variants.

The N=1 Collaborative (N1C) (n1collaborative.org/) is a global initiative to standardize ultra-rare “n-of-1/few” therapy development for safe and equitable delivery to individuals with rare diseases. The N1C VARIANT (**V**ariant **A**ssessments towa**r**ds El**i**gibility for **An**tisense Oligonucleotide **T**reatment) guidelines were recently developed to standardize first steps in the complex process of adjudicating DNA variants for eligibility to existing ASO mechanisms of action.^11^ The goal of this study was to screen the eligibility status of all diagnoses made by clinical GWS in a large pediatric hospital over a six-year period.

## MATERIALS AND METHODS

### Cohort definition

This retrospective cohort study was approved by the SickKids Research Ethics Board (#1000079841), with an accompanying patient consent waiver and with only public DNA variants described in detail. All information used for assessments was from electronic medical records. Phenotypic information was codified using the Human Phenotype Ontology (HPO). Probands with a partial or complete genetic diagnosis, via clinical GWS resulted between 01-Jul-2018 and 30-Jun-2024, were included in the study. GWS results were considered diagnostic if: (i) the variant(s) was/were classified as likely pathogenic or pathogenic according to ACMG/AMP guidelines, with the necessary zygosity to be disease-causing, and (ii) available medical records indicated that the variant(s) was/were considered by the clinical team as explanatory for some or all the primary phenotype(s). Indications for ordering GWS in the province of Ontario during this study period are described elsewhere.^2^

### Variant assessment for ASO amenability

We applied version 1.0 of the N1C VARIANT guidelines.^11^ The primary ASO approaches considered were canonical exon skipping, splice correction, and mRNA knockdown. Variants were classified as either “eligible”, “likely eligible”, “unlikely eligible”, or “not eligible” in relation to the different ASO approaches, or “unable to assess”. All patient variants were initially assessed by a single individual (author D.C.). A subset of variants was reviewed by N1C Patient Identification Working Group members (authors M.L., L.N.) blind to the initial classification, to confirm correct application of the VARIANT guidelines. The evidence supporting an “eligible” or “likely eligible” designation for each variant, and other disease/clinical considerations, were discussed by the SickKids Advanced Therapeutics Triage Working Group (authors K.A., M.A., A.R.D., J.J.D., Z.I., R.O., A.P., M.R.). Genes with a loss-of-function allele in trans to a wild-type allele were also analyzed to determine whether they included poison exons, naturally occurring antisense transcripts, and/or upstream open reading frames.^11^

## RESULTS

### Cohort demographics and variant characteristics

Of the 532 consecutive patients with molecular genetic diagnoses (55% male), the median number of positive HPO indications for testing was 9 (range 1-43) (Supplemental Table 2). The most common testing strategies were trio exome sequencing (65%) and trio genome sequencing (21%) (Supplemental Figure 1). Median (mean) age at time of positive GWS result was 3 years (5.5 years). There were 592 total diagnostic variants across 449 different disease genes. Most variants were missense variants (Supplemental Table 3). Most genes were associated with neurological and/or neurodevelopmental disorders (Supplemental Figure 2), and 39% of genes were previously curated by the Dutch Centre for RNA Therapeutics^6^ with respect to disease-related treatment candidacy.

### Variant eligibility for canonical exon skipping

We assessed 519 variants that were contained within a single exon for exon skipping, irrespective of pathomechanism. Exons were ineligible for potential skipping if, e.g., they are the first or last coding exon of the gene, a deletion would result in a frameshift, nested in-frame deletions are pathogenic, and/or they contain key functional protein domains or mutational hotspot (Supplemental Figure 3).^11^ Four variants (0.68%) were identified as “likely eligible” [*ARID2*(NM_152641.2) exon 10, *PCNT*(NM_006031.6) exon 34, *SMPD4*(NM_017951.4) exon 10, and *VPS13B*(NM_017890.3) exon 9)] (Table 1; Supplemental Table 4; Supplemental Figure 3). Fourteen additional variants (2.36%) were classified as “unlikely eligible” because of size or protein domain considerations (Supplemental Table 4; Supplemental Figure 3). None of the 12 single or multi-exon deletions impacted genes in which skipping an additional exon was expected to restore reading frame and protein function.

### Variant eligibility for splice correction

We considered intronic and exonic variants separately. First, 48 variants from our patient cohort are intronic or affect the intron-exon boundary. The VARIANT guidelines^11^ require functional evidence to properly assess a variant towards splice correction (i.e., *in silico* splicing predictors are not suitable substitutes),^12,13^ and functional evidence was only publicly available for 8 variants (Supplemental Table 5). Regardless, even if functional evidence were available for all variants, 42 are within 5 base pairs of the intron-exon border and are therefore classified as “not eligible” towards splice correction ASOs.^11^ The 6 remaining intronic variants were NM_001089.3*(ABCA3*):c.3863-98C>T (ClinVar VCV001211382.11), NM_003748.4(*ALDH4A1*):c.679-6G>A (ClinVar VCV001007564.7), NM_004380.3(*CREBBP*):c.2881-13G>A (ClinVar VCV001397911.6), NM_000518.5 (*HBB*):c.316-106C>G (ClinVar VCV000015457.121), and 2 previously unreported variants (a 12 bp deletion impacting the donor site of intron 1 of NM_001399.5(*EDA*) and a SNV near the acceptor site of exon 19 of NM_004247.4(*EFTUD2*). The *ALDH4A1, CREBBP, EDA*, and *EFTUD2* variants would be considered “unlikely eligible” based on position (i.e., likely impacting branchpoint or donor splicing motifs), if functional evidence of aberrant splicing were available. The deep intronic variants in *ABCA3* and *HBB* have proven mis-splicing effects (Supplemental Figure 4), and were classified as “likely eligible” (Table 1).

An additional 3 synonymous exonic variants were known experimentally to result in mis-splicing: NM_001110556.2(*FLNA*):c.1923C>T (ClinVar VCV000011770.29), NM_001110556.2(*FLNA*):c.5217G>A (ClinVar VCV000011775.40), and NM_024649.5(*BBS1*):c.1110G>A (ClinVar VCV000553357.14). The first variant in *FLNA* creates a new donor site 93 bases upstream, resulting in a 101bp deletion from the 3’ end of exon 13,^14^ and is classified as “likely eligible” (Supplemental Figure 5). The latter two variants are “not eligible” since they are within 5 base pairs of the exon-intron boundary.

### Variant eligibility for mRNA knockdown

From our cohort, 31 variants had robust evidence for either a toxic gain-of-function or dominant-negative effect (Supplemental Table 6). Two were classified as “eligible”: *PACS1* variant based on pre-clinical evidence^15^ and a report of work in progress via the n-Lorem Foundation^10^,and *MYH7* variant based on pre-clinical evidence. ^16^ An additional 16 were “likely eligible” because haploinsufficiency was tolerated, and 13 were “unlikely eligible” because haploinsufficiency is associated with disease (Table 1). There were also 90 variants across 65 genes classified as “unable to assess” due to lack of experimental confirmation of variant pathomechanism, despite reports of different gain-of-function or dominant-negative pathogenic variants in these genes (Supplemental Table 7). If these 90 variants were confirmed to have gain-of-function or dominant-negative effects, 5 would be considered “eligible”, 24 “likely eligible”, and 61 “unlikely eligible” (Supplemental Table 7).

### Clinical and feasibility considerations regarding targeting identified variants with ASOs

Overall, 26 of 592 diagnostic variants (4.4%; in n=26 different patients) were assessed as “eligible” or “likely eligible” to ASO correction (via canonical exon skipping, splice correction, or mRNA knockdown) based on variant-level factors and stringent criteria (Table 1). Clinical and demographic features of these twenty-six individuals were similar to the remainder of the cohort (Supplemental Table 2). Only eight diagnoses were made within a year of symptom onset.

The VARIANT guidelines do not consider tissue targetability or other clinical considerations when classifying variants.^11^ After considering disease and delivery factors,^4,5^ 11 of 26 diagnoses were still considered candidates for bespoke ASO development (Supplemental Table 8). For example, the variant in *ABCA3* was “likely eligible” but efficient administration of ASOs to pulmonary tissue is not yet possible.

### Variant eligibility for a wildtype allele upregulation approach

Wildtype allele upregulation is a promising but relatively less established approach for ASO therapies;^5,17^ designating eligibility is not yet codified in the VARIANT guidelines.^11^ We assessed the future potential for wildtype allele upregulation in this study cohort by considering two necessary (but not sufficient) criteria for this approach. First, there were 234 variants in 144 different disease genes that were thought to result in loss-of-function in trans to a wildtype allele.

Second, poison exons, naturally occurring antisense transcripts, and/or upstream open reading frames were listed in at least 1 of 5 publications/databases for 221 of these variants; Supplemental Figure 6).

## DISCUSSION

We screened the diagnoses made by clinical GWS in 532 consecutive individuals at a single centre for ASO amenability, using the N1C VARIANT guidelines. The results suggest that a meaningful proportion of the diagnostic variants could be targets for ASO treatment using current established methods. Considering every molecular genetic diagnosis for genetic therapy eligibility parallels growing interest in expanding the “interventionalist” role within clinical genetics.^18^ Our findings also underscore the importance of timely genetic diagnosis and of ancillary testing to clarify pathomechanism, which remain priority issues for the broader medical genetics community.

High-profile proofs-of-concept are generating questions in the rare disease community about the potential for individualized ASO development for specific variants. Applying the N1C VARIANT guidelines to a real-world cohort may help to demystify why some rare genetic diagnoses are deemed more tractable to current ASO methods than others.^7^ No evaluation approach will have complete sensitivity and specificity, and variant assessment is only one component of the complex ASO evaluation and development process. Our approach is on the spectrum in between methods used in two prior reports,^10,17^ and has the advantage of involving a more representative clinical cohort. One report assessed pathogenic variants in ClinVar for potential amenability to wildtype upregulation approaches and suggested an upper bound of 50% for ASO amenability.^17^ The other report looked at outcomes from review by expert committee, for 173 individuals with nano-rare variants from across the United States and Canada nominated for the n-Lorem Foundation ASO development program over a multi-year period.^10^ Developing, debating, applying, and refining the VARIANT guidelines will support broader access to “variant triage” and efforts to scale to a population level.

There are additional limitations of our study. Assessment of ASO amenability can change over time, including because of advancements in technology. We did not consider genetic diagnoses made by more targeted genetic testing (e.g., next-generation sequencing gene panels). We did not have access to patient-derived cells or other model systems to develop and test ASOs,^19^ nor the opportunity to administer ASOs and determine clinical efficacy. False negative and false positive screening rates cannot yet be calculated. We did not interview these patients, families, or their clinicians to assess their overall interest, and perception of risks and benefits, of an ASO therapy. Last, we acknowledge the clinical and ethical dimensions of decision-making regarding resource allocation in this individualized genetic therapy space, which were beyond the scope of this report.

In summary, our findings support the need for proactive identification and accelerated testing of genetic variants amenable to ASO treatments. Balancing expert guidance/oversight with increasing global demand for variant screening will continue to be important. However, relieving some of the pressure on individuals and families to self-advocate by increasing awareness of ASO eligibility considerations among clinicians could reduce health inequities. How best to share variant annotations for ASO amenability with the larger genetics community warrants further discussion, and could leverage successful existing databases like ClinVar. In time, assessments may also expand to other bespoke genetic therapy approaches.^20^

## FIGURE LEGEND

**Figure 1.**
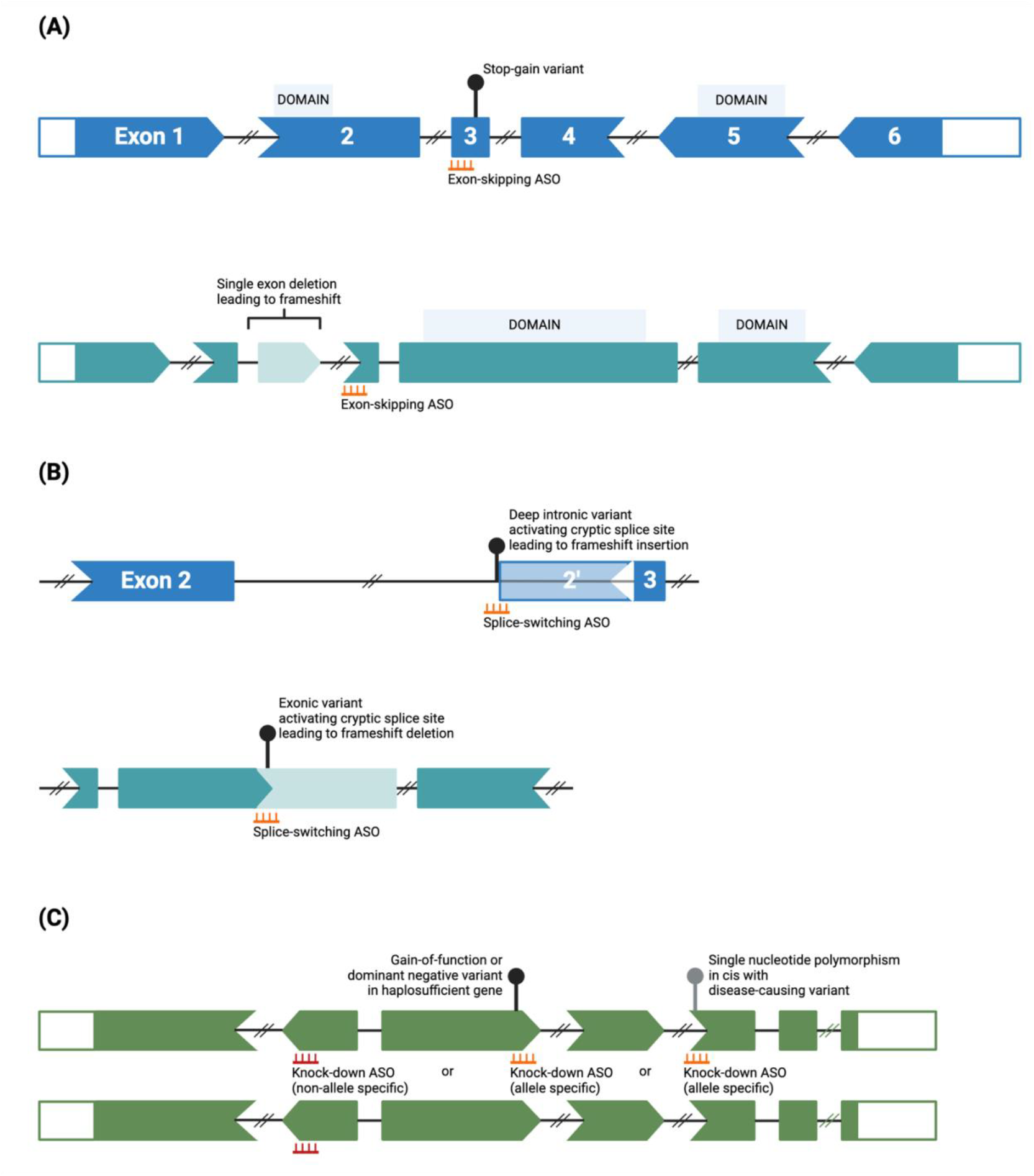
Overview of ASO strategies considered in the N1C VARIANT guidelines. Created in BioRender. (A)Canonical exon skipping ASOs can be used to skip an exon, with the goal of producing a truncated yet functional protein product. An in-frame exon containing, e.g., a premature stop codon can be skipped (top transcript), or an exon can be skipped to restore the reading frame (bottom transcript). (B)Splice correcting ASOs can be used to restore canonical splicing when the variant pathomechanism is aberrant splicing. ASOs can target and mask intronic (top) or exonic (bottom) cryptic splice sites. (C)ASOs can be used to knockdown alleles containing dominant negative or gain of function variants. ASOs can be allele specific (orange) or non-allele specific (red). Allele-specific ASOs can target either single-nucleotide polymorphisms in cis with the variant, or the variant itself (top).

## Data Availability

All data produced in the present work are contained in the manuscript

## Acknowledgements

We thank the children and their families who formed the study cohort, and the many clinicians involved in their diagnosis and care. Special thanks to the Genome-wide Sequencing Ontario program, the SickKids DNA Resource Centre, and SickKids Precision Child Health including Translational Genomics and Advanced Therapeutics team members. D.C. is supported by a SickKids Restracomp Master's Scholarship and the SickKids Innovators Fund. M.C.L is funded by a Walter-Benjamin Fellowship by the German Research Foundation. D.B. is supported by a Humboldt Research Fellowship for Postdocs and the Hertie-Network of Excellence in Clinical Neuroscience. A.R.D is funded by the Azrieli Precision Child Health Platform. G.C. acknowledges support from the University of Toronto McLaughlin Centre and Canadian Institutes of Health Research (PJT186240).

## REFERENCES

1. Howell KB, White SM, McTague A, et al. International Precision Child Health Partnership (IPCHiP): an initiative to accelerate discovery and improve outcomes in rare pediatric disease. NPJ Genom Med. 2025;In press.

2. Haque B, Khan T, Ushcatz I, et al. Contemporary aetiologies of medical complexity in children: a cohort study. Arch Dis Child. 2023;108(2):147–149.

3. Boycott KM, Hartley T, Biesecker LG, et al. A Diagnosis for All Rare Genetic Diseases: The Horizon and the Next Frontiers. Cell. 2019;177(1):32–37.

4. Roberts TC, Langer R, Wood MJA. Advances in oligonucleotide drug delivery. Nat Rev Drug Discov. 2020;19(10):673–694.

5. Lauffer MC, van Roon-Mom W, Aartsma-Rus A, Collaborative N. Possibilities and limitations of antisense oligonucleotide therapies for the treatment of monogenic disorders. Commun Med (Lond). 2024;4(1):6.

6. Aartsma-Rus A, Collin RWJ, Elgersma Y, Lauffer MC, van Roon-Mom W. Joining forces to develop individualized antisense oligonucleotides for patients with brain or eye diseases: the example of the Dutch Center for RNA Therapeutics. Ther Adv Rare Dis. 2024;5:26330040241273465.

7. Kim J, Woo S, de Gusmao CM, et al. A framework for individualized splice-switching oligonucleotide therapy. Nature. 2023;619(7971):828–836.

8. Kim J, Hu C, Moufawad El Achkar C, et al. Patient-Customized Oligonucleotide Therapy for a Rare Genetic Disease. N Engl J Med. 2019;381(17):1644–1652.

9. Ziegler A, Carroll J, Bain JM, et al. Antisense oligonucleotide therapy in an individual with KIF1A-associated neurological disorder. Nat Med. 2024;30(10):2782–2786.

10. Crooke ST, Cole T, Carroll JB, et al. Genotypic and phenotypic analysis of 173 patients with extremely rare pathogenic mutations who applied for experimental antisense oligonucleotide treatment. medRxiv. 2024:2024.2008.2005.24310862.

11. Cheerie D, Meserve M, Beijer D, et al. Consensus guidelines for eligibility assessment of pathogenic variants to antisense oligonucleotide treatments. medRxiv. 2024:2024.2009.2027.24314122.

12. Oh RY, AlMail A, Cheerie D, et al. A systematic assessment of the impact of rare canonical splice site variants on splicing using functional and in silico methods. HGG Adv. 2024;5(3):100299.

13. Deshwar AR, Yuki KE, Hou H, et al. Trio RNA sequencing in a cohort of medically complex children. Am J Hum Genet. 2023;110(5):895–900.

14. Hehr U, Hehr A, Uyanik G, Phelan E, Winkler J, Reardon W. A filamin A splice mutation resulting in a syndrome of facial dysmorphism, periventricular nodular heterotopia, and severe constipation reminiscent of cerebro-fronto-facial syndrome. J Med Genet. 2006;43(6):541–544.

15. Villar-Pazos S, Thomas L, Yang Y, et al. Neural deficits in a mouse model of PACS1 syndrome are corrected with PACS1- or HDAC6-targeting therapy. Nat Commun. 2023;14(1):6547.

16. Dainis A, Zaleta-Rivera K, Ribeiro A, et al. Silencing of MYH7 ameliorates disease phenotypes in human iPSC-cardiomyocytes. Physiol Genomics. 2020;52(7):293–303.

17. Mittal S, Tang I, Gleeson JG. Evaluating human mutation databases for “treatability” using patient-customized therapy. Med. 2022;3(11):740–759.

18. Pena LDM, Burrage LC, Enns GM, et al. Contributions from medical geneticists in clinical trials of genetic therapies: A points to consider statement of the American College of Medical Genetics and Genomics (ACMG). Genet Med. 2023;25(6):100831.

19. Means JC, Martinez-Bengochea AL, Louiselle DA, et al. Rapid and scalable personalized ASO screening in patient-derived organoids. Nature. 2025.

20. Dowling JJ, Pirovolakis T, Devakandan K, et al. AAV gene therapy for hereditary spastic paraplegia type 50: a phase 1 trial in a single patient. Nat Med. 2024;30(7):1882–1887.

