## Supplemental File for "Screening rare genetic diagnoses for amenability to bespoke antisense oligonucleotide therapy development: a retrospective cohort study"

Supplemental Table 1. List of FDA-approved oligonucleotide therapies for the treatment of genetic conditions. 2

Supplemental Table 2. Features of the study cohort, including the subgroups with and without diagnostic variants considered "eligible"/"likely eligible" for ASO therapies. 3

Supplemental Table 3. Categories of diagnostic variants (n=592) in the study cohort. 4

Supplemental Table 4. Details of "likely eligible" and "unlikely eligible" variants for exon skipping. 5

Supplemental Table 5. Assessment of functionally validated intronic splicing variants for splice correcting ASOs. 7

Supplemental Table 6. Assessment of "eligible", "likely eligible" and "unlikely eligible" exons for RNA knockdown. 8

Supplemental Table 7. Variants with unconfirmed pathomechanisms identified in genes where GoF and/or DN-acting variants are possibly associated with disease. 10

Supplemental Table 8. Disease/condition- and delivery-related considerations for eligible and likely eligible variants identified in this study. 14

Supplemental Figure 1. Clinical genome-wide sequencing tests and family designs used in the diagnosis of individuals in this study cohort. 18

Supplemental Figure 2. Disease category associations for genes (n=449) containing diagnostic variants in the study cohort. 19

Supplemental Figure 3. Overview of exon skipping assessment for diagnostic variants in the study cohort. 20

Supplemental Figure 4. Aberrant splicing caused by *ABCA3* and *HBB* variants in the study cohort. 21

Supplemental Figure 5. Aberrant splicing caused by an *FLNA* variant in the study cohort. 22

Supplemental Figure 6. Number of variants in the study cohort for which the corresponding genes may have poison exons, naturally occurring antisense transcripts, and/or upstream open reading frames. 23

**Supplemental Table 1. List of FDA-approved oligonucleotide therapies for the treatment of genetic conditions.**

| Drug name (year of approval) | Category (mode of action) | Target gene | Condition |
|---|---|---|---|
| Givosiran (2019) | siRNA (RISC) | *ALAS1* | Acute hepatic porphyria |
| Mipomersen (2013) | ASO (RNAse H1) | *APOB* | Familial hypercholesterolemia |
| Eteplirsen (2016) | ASO (exon skipping) | *DMD* | Duchenne muscular dystrophy |
| Golodirsen (2019) | ASO (exon skipping) | *DMD* | Duchenne muscular dystrophy |
| Viltolarsen (2020) | ASO (exon skipping) | *DMD* | Duchenne muscular dystrophy |
| Casimersen (2021) | ASO (exon skipping) | *DMD* | Duchenne muscular dystrophy |
| Lumasiran (2020) | siRNA (RISC) | *HOA1* | Primary hyperoxaluria type I |
| Nedosiran (2023) | siRNA (RISC) | *HOA1* | Primary hyperoxaluria type I |
| Inclisiran (2021) | siRNA (RISC) | *PCSK9* | Primary hyperlipidemia |
| Nusinersen (2016) | ASO (exon inclusion) | *SMN2* | Spinal muscular atrophy |
| Tofersen (2023) | ASO (RNAse H1) | *SOD1* | SOD1-associated ALS |
| Patisiran (2018) | siRNA (RISC) | *TTR* | ATTRv amyloidosis |
| Inotersen (2018) | ASO (RNAse H1) | *TTR* | ATTRv amyloidosis |
| Vutrisiran (2022) | siRNA (RISC) | *TTR* | ATTRv amyloidosis |
| Eplontersen (2023) | ASO (RNAse H1) | *TTR* | Transthyretin mediated amyloid cardiomyopathy |

Adapted from [1] and updated by searching drugbank.com in January 2025.

ALS, amyotrophic lateral sclerosis; ASO, antisense oligonucleotide; ATTRv amyloidosis, hereditary transthyretin amyloidosis; FDA, Food and Drug Administration (United States); RISC, RNA-induced silencing complex; RNAse, ribonuclease; siRNA, small interfering RNA

**Supplemental Table 2. Features of the study cohort, including the subgroups with and without diagnostic variants considered "eligible"/"likely eligible" for ASO therapies.**

| Feature | Study cohort (n=532) | Patients with eligible or likely eligible variants (n=26) | Patients without eligible or likely eligible variants (n=506) |
|---|---|---|---|
| Average age in years | 5.5 | 5.3 | 5.5 |
| Median age in years (range) | 3 (0-25) | 4 (0-18) | 3 (0-25) |
| Male sex | 55% | 54% | 55% |
| Median HPO terms (range) | 9 (1-43) | 9 (5-35) | 9 (1-43) |
| Selected HPO terms: | | | |
| Developmental delay | 300 (56%) | 17 (65%) | 283 (56%) |
| Hypotonia | 181 (34%) | 9 (35%) | 172 (34%) |
| Seizures | 109 (20%) | 6 (23%) | 103 (20%) |
| Microcephaly | 94 (18%) | 5 (19%) | 89 (18%) |
| Failure to thrive[a] | 88 (17%) | 3 (12%) | 85 (17%) |
| Exome sequencing (%)[b] | 419 (79%) | 20 (77%) | 399 (79%) |
| Total number of diagnostic variants (unique variants) | 592 (588) | 26 (25) | 566 (563) |
| Unique disease genes | 449 | 24 | 432 |
| Genes curated by DCRT[c] | 173 | 9 | 169 |
| DCRT eligible genes[d] (% of curated) | 131 (76%) | 7 (78%) | 129 (76%) |

DCRT, Dutch Centre for RNA Therapeutics; HPO, Human Phenotype Ontology

[a]"Failure to thrive" is currently the HPO term but in our region this terminology is now considered outdated and potentially stigmatizing.
[b]The remainder had short-read genome sequencing as a clinical genome-wide sequencing test.
[c]See Aartsma-Rus *et al.* (2024) for details.[2]
[d]Genes associated with "progressive neurological disorder with predominant CNS phenotype" or "slowly progressive neurological diseases, diseases with questionable progression, and progressive diseases with extra-neuronal manifestation that could still be eligible for treatment".

**Supplemental Table 3. Categories of diagnostic variants (n=592) in the study cohort.**

| Variant category | Number of variants | % |
|---|---:|---:|
| Missense single nucleotide variant[a] | 251 | 42.4% |
| Frameshift variant (within an exon)[a] | 138 | 23.3% |
| Nonsense single nucleotide variant[a] | 100 | 16.9% |
| Intronic single nucleotide variant[b] | 42 | 7.1% |
| In-frame deletion (within an exon)[a] | 17 | 2.9% |
| Partial gene (whole or multi-exon) deletion | 12 | 2.0% |
| Multigene deletion | 7 | 1.2% |
| Multigene duplication | 5 | 0.8% |
| Synonymous single nucleotide variant[a] | 5 | 0.8% |
| In-frame duplication (within an exon)[a] | 4 | 0.7% |
| Insertion and deletion (within an exon)[a] | 3 | 0.5% |
| Deletion (within an intron) | 2 | 0.3% |
| Exon-intron boundary deletion | 2 | 0.3% |
| Insertion and deletion (within an intron) | 1 | 0.2% |
| Duplication (within an intron) | 1 | 0.2% |
| Partial gene (multi-exon) duplication | 1 | 0.2% |
| Deletion of canonical start-site[a] | 1 | 0.2% |

[a]Considered for exon skipping in Supplemental Figure 3.
[b]Including n=31 canonical splice site variants, n=9 variants at distance 3-15 bp from canonical splice sites, and n=2 deep intronic variants.

**Supplemental Table 4. Details of "likely eligible" and "unlikely eligible" variants for exon skipping.**

See text for details. No variants in the study cohort were designated "eligible". Reasons for variants being "not eligible" are depicted in Supplemental Figure 3.

| Gene symbol (MIM #) | Transcript | Variant type | Exon | N1C VARIANT classification | Reasoning |
|---|---|---|---|---|---|
| *ARID2* (609539) | NM_152641.2 | Frameshift | 10 | Likely | • Small exon (<10% of total cDNA length)<br>• In-frame exon<br>• Not known to code for a functional domain<br>• No missense variants or in-frame deletions within the exon reported as a cause of disease |
| *PCNT* (605925) | NM_006031.6 | Frameshift | 34 | | |
| *SMPD4* (610457) | NM_017951.4 | Frameshift | 10 | | |
| *VPS13B* (607817) | NM_017890.3 | Frameshift | 9 | | |
| *ACTL6B* (612458) | NM_016188.4 | Missense | 12 | Unlikely | Small, in-frame exon, but the pathogenicity of this homozygous missense variant may imply an important functional role encoded by exon 12 |
| *AP3B1* (603401) | NM_003664.5 | Frameshift | 4 | | Small, in-frame exon with no reported pathogenic missense variants. Exon encodes part of a conserved N-terminal domain, though its role has not been functionally proven |
| *CDK13* (603309) | NM_003718.5 | Nonsense | 2 | | Large domain that encodes a serine-arginine rich region, though its functional role has not been experimentally validated |
| *CHD2* (602119) | NM_001271.4 | Frameshift | 7 | | Small, in-frame exon with no reported pathogenic missense variants or in-frame deletions. Exon codes for disordered region and has compositional bias. |
| *COL4A1* (120130) | NM_001845.6 | Missense | 11 | | Small, in-frame exon, with reported missense variants effecting the glycine in the important G-X-Y repeats (such as our patient's variant). Exon encodes these G-X-Y repeats, and is therefore unlikely according to our guidelines |
| *CREBBP* (600140) | NM_004380.3 | Nonsense | 14 | | Small, in-frame exon with no reported pathogenic missense or in-frame variants. Exon codes for a linker between two domains. |
| *EP300* (602700) | NM_001429.4 | Nonsense | 13 | | Small, in-frame exon with no reported pathogenic missense or in-frame variants. Exon codes for a disordered region. |
| *GLDN* (608603) | NM_181789.4 | Nonsense | 6 | | Small, in-frame exon with no reported pathogenic missense or in-frame |

| | | | | | |
|---|---|---|---|---|---|
| | | | | | variants. Exon codes for part of a topological domain with unspecified function. |
| *JAG1* (601920) | NM_000214.3 | Nonsense | 21 | | Small, in-frame exon with no reported pathogenic missense or in-frame variants. Exon within a repeat domain. |
| *KAT6A* (601408) | NM_006766.5 | Nonsense | 15 | | In-frame exon coding for more than 10% of the coding transcript. Exon codes for a compositional bias (acidic domain). |
| *KATNIP* (616650) | NM_015202.5 | Nonsense | 16 | | In-frame exon coding for more than 10% of the coding transcript. Exon codes for a domain of unknown function. No reported pathogenic missense or in-frame variants. |
| *SETD5* (615743) | NM_001080517.3 | Nonsense | 19 | | In-frame exon coding for more than 10% of the coding transcript. No reported pathogenic missense or in-frame variants. Exon codes for a disordered region. |
| *TRRAP* (603015) | NM_003496.3 | Missense | 26 | | Small, in-frame exon with no reported pathogenic missense or in-frame variants. (apart from this individual's missense variant). Within a repeat domain. |
| *ZMIZ1* (607159) | NM_020338.4 | Missense | 14 | | Small, in frame exon for which skipping has been observed in both healthy and non-healthy individuals. No functional studies exist on the role of exon 14 skipping, and our patient has a missense variant. |

**Supplemental Table 5. Assessment of functionally validated intronic splicing variants for splice correcting ASOs.**

<table>
<tr><th>DNA variant</th><th>ClinVar accession number</th><th>N1C VARIANT classification</th><th>Reasoning</th></tr>
<tr><td>NM_001089.2(ABCA3):c.3863-98C>T</td><td>VCV001211382.11</td><td rowspan="2">Likely eligible</td><td rowspan="2">Deep intronic variant resulting in formation of a pseudoexon; canonical splice sites and branchpoint not weakened or destroyed</td></tr>
<tr><td>NM_000518.5(HBB):c.316-106C>G</td><td>VCV000015457.121</td></tr>
<tr><td>NM_012079.5(DGAT):c.751+2T>C</td><td>VCV000139512.20</td><td rowspan="6">Not eligible</td><td rowspan="6">Within 5bp of the canonical splice site; canonical splice site is destroyed</td></tr>
<tr><td>NM_000515.3(GH1):c.291+5G>A</td><td>VCV000015975.2</td></tr>
<tr><td>NM_000516.7(GNAS):c.432+1G>A</td><td>VCV000446491.12</td></tr>
<tr><td>NM_001271043.2(NFIX):c.979+1G>A</td><td>VCV000036964.4</td></tr>
<tr><td>NM_004278.3(PIGL):c.336-2A>G</td><td>VCV000632268.8</td></tr>
<tr><td>NM_001292034.3(TAB2):c.1764+1G>A</td><td>VCV001030404.8</td></tr>
</table>

**Supplemental Table 6. Assessment of "eligible", "likely eligible" and "unlikely eligible" exons for RNA knockdown.**

| DNA variant | ClinVar accession number | N1C VARIANT classification | Reasoning |
|---|---|---|---|
| NM_018026.3(*PACS1*):c.607C>T | VCV000039581.95 | Eligible | Evidence of knockdown ASO in pre-clinical studies |
| NM_000257.4(*MYH7*):c.2155C>T | VCV000014104.38 | | |
| NM_172362.2(*KCNH1*):p.(G496E) | [PMID: 26818738] | Likely eligible | Haploinsufficiency not associated with disease, to the best of our knowledge |
| NM_012062.5(*DNM1L*): c.1207C>T | VCV000214313.49 | | |
| NM_000142.5(*FGFR3*):c.1138G>A | VCV000016328.58 | | |
| NM_000142.5(*FGFR3*):c.1172C>A | VCV000016329.30 | | |
| NM_006086.4(*TUBB3*):c.904G>A | VCV000006964.28 | | |
| NM_014225.6(*PPP2R1A*):c.656C>T[a] | VCV000521503.43 | | |
| NM_001303256.2(*MORC2*):c.79G>A | VCV000804164.25 | | |
| NM_004333.4(*BRAF*):c.1785T>G | VCV000177672.14 | | |
| NM_005052.3(*RAC3*):c.186_188del | VCV001723153.1 | | |
| NM_001927.4(*DES*):c.1216C>T | VCV000016826.29 | | |
| NM_002730.4 (*PRKACA*):c.409G>A | VCV000989460.5 | | |
| NM_003289.4(*TPM2*):c.415_417del | VCV000012465.26 | | |
| NM_000515.3(*GH1*):c.291+5G>A | VCV000015975.2 | | |
| NM_000352.6(*ABCC8*):c.4613G>A | VCV000585348.23 | | |
| NM_006245.3(*PPP2R5D*):c.592G>A | VCV000190286.64 | | |
| NM_005188.4(*CBL*):c.1150T>C | VCV000029822.8 | | |
| NM_172107.4(*KCNQ2*):c.430C>T | VCV000452487.38 | Unlikely eligible | Haploinsufficiency reported to be a cause of disease[c] |
| NM_000217.3(*KCNA1*):c.1221T>G | [PMID: 23349320] | | |

| | |
|---|---|
| NM_000828.4(*GRIA3*):*c.2101A>G*[b] | [PMID: 38038360] |
| NM_002074.4(*GNB1*):c.233A>G | VCV000224714.4 |
| NM_000806.5(*GABRA1*):c.875C>G | VCV003253005.1 |
| NM_002834.5(*PTPN11*):c.1403C>T | VCV000013331.86 |
| NM_002834.5(*PTPN11*):c.794G>A | VCV000040522.68 |
| NM_001127221.1(*CACNA1A*)c.2137G>A | VCV000254268.47 |
| NM_014233.4(*UBTF*):c.628G>A | VCV000437909.31 |
| NM_000552.5(*VWF*):c.3614G>T | VCV000100271.2 |
| NM_001320.7(*CSNK2B*):c.94G>A | VCV000520596.14 |
| NM_003924.4(*PHOX2B*):c.738_776dup | VCV000984923.4 |
| NM_024665.7(*TBL1XR1*):c.1337A>G | VCV000225874.10 |

[a]Variant found in two different individuals in the study cohort.
[b]X inactivation possible, therefore knockdown may not be a suitable approach
[c]The N1C VARIANT guidelines version 1.0 do not factor in the nature of the disease caused by haploinsufficiency (e.g., whether it is markedly less severe than the disease for which ASO knockdown of a variant is being considered). Also, for genes where there was provisional or conflicting evidence for haploinsufficiency being associated with disease, we conservatively considered variants in these genes to be “unlikely eligible”.

**Supplemental Table 7. Variants with unconfirmed pathomechanisms identified in genes where GoF and/or DN-acting variants are possibly associated with disease.**

| # of DNA variants | Gene | OMIM # | N1C VARIANT classification | Reasoning |
|---|---|---|---|---|
| 3 | *ACTA1* | 102610 | Likely eligible | Missense variant in gene where GoF or DN mechanism are cause of disease, but not functionally proven for specific variant(s) in our cohort |
| 2 | *ACTB* | 102630 | Likely eligible | Missense variant in gene where GoF or DN mechanism are cause of disease, but not functionally proven for specific variant(s) in our cohort |
| 1 | *ACTG2* | 102545 | Likely eligible | Missense variant in gene where GoF or DN mechanism are cause of disease, but not functionally proven for specific variant(s) in our cohort |
| 1 | *ADNP* | 615873 | Unlikely eligible | Truncating variant in last exon where LoF or GoF/DN mechanisms are possible |
| 1 | *AFG3L2* | 604581 | Likely eligible | Missense variant in gene where GoF or DN mechanism are cause of disease, but not functionally proven for specific variant(s) in our cohort |
| 2 | *AHDC1* | 615829 | Unlikely eligible | Truncating variant in last exon where LoF or GoF/DN mechanisms are possible |
| 1 | *ATP1A3* | 182350 | Unlikely eligible | Missense variant in gene where LoF or GoF/DN mechanisms are possible |
| 1 | *ATP6V1A* | 607027 | Unlikely eligible | Missense variant in gene where LoF or GoF/DN mechanisms are possible |
| 1 | *BRAF* | 164757 | Likely eligible | Missense variant in gene where GoF or DN mechanism are cause of disease, but not functionally proven for specific variant(s) in our cohort |
| 4 | *CACNA1A* | 601011 | Unlikely eligible | Missense or in-frame deletion variants in gene where LoF or GoF/DN mechanisms are possible |
| 2 | *CAMK2B* | 607707 | Unlikely eligible | Missense variant in gene where LoF or GoF/DN mechanisms are possible |
| 1 | *CDK13* | 603309 | Unlikely eligible | Missense variant in gene where LoF or GoF/DN mechanisms are possible |
| 1 | *CHD3* | 602120 | Unlikely eligible | Missense variant in gene where LoF or GoF/DN mechanisms are possible |
| 1 | *CHD7* | 608892 | Unlikely eligible | Missense variant in gene where LoF or GoF/DN mechanisms are possible |
| 1 | *CHN1* | 118423 | Likely eligible | Missense variant in gene where GoF or DN mechanism are cause of disease, but not functionally proven for specific variant(s) in our cohort |
| 2 | *COL4A1* | 120130 | Unlikely eligible | Missense variant in gene where LoF or GoF/DN mechanisms are possible |
| *2* | *DDX3X* | 300160 | Unlikely eligible | Missense variant in gene where LoF or GoF/DN mechanisms are possible |
| 2 | *DYNC1H1* | 600112 | Likely eligible | Missense variant in gene where GoF or DN mechanism are cause of disease, but not functionally proven for specific variant(s) in our cohort |
| 1 | *DYRK1A* | 600855 | Unlikely eligible | Missense variant in gene where LoF or GoF/DN mechanisms are possible |

| | | | | |
|---|---|---|---|---|
| 1 | *EEF1A2* | 602959 | Likely eligible | Missense variant in gene where GoF or DN mechanism are cause of disease, but not functionally proven for specific variant(s) in our cohort |
| 2 | *ERF* | 611888 | Unlikely eligible | Missense variant in gene where LoF or GoF/DN mechanisms are possible |
| 1 | *FOXF1* | 601089 | Unlikely eligible | Missense variant in gene where LoF or GoF/DN mechanisms are possible |
| 1 | *FOXJ1* | 602291 | Unlikely eligible | Truncating variant in the last exon, in a gene where LoF or GoF/DN mechanisms are possible |
| 1 | *FBXW7* | 606278 | Unlikely eligible | Missense variant in gene where LoF or GoF/DN mechanisms are possible |
| 1 | *FLNA* | 300017 | Unlikely eligible | Missense variant in gene where LoF or GoF/DN mechanisms are possible |
| 1 | *GABBR2* | 607340 | Unlikely eligible | Missense variant in gene where LoF or GoF/DN mechanisms are possible |
| 1 | *GABRA5* | 137142 | Unlikely eligible | Missense variant in gene where LoF or GoF/DN mechanisms are possible |
| 1 | *GRIN1* | 138249 | Unlikely eligible | Missense variant in gene where LoF or GoF/DN mechanisms are possible |
| 1 | *GRIN2A* | 138253 | Unlikely eligible | Missense variant in gene where LoF or GoF/DN mechanisms are possible |
| *2* | *GRIN2D* | 602717 | Unlikely eligible | Missense variant in gene where LoF or GoF/DN mechanisms are possible |
| 1 | *HUWE1* | 300697 | Unlikely eligible | Missense variant in gene where LoF or GoF/DN mechanisms are possible |
| 2 | *KCNB1* | 600397 | Unlikely eligible | Missense variant in gene where LoF or GoF/DN mechanisms are possible |
| 1 | *KCND2* | 605410 | Unlikely eligible | Missense variant in gene where LoF or GoF/DN mechanisms are possible |
| 1 | *KCND3* | 605411 | Unlikely eligible | Missense variant in gene where LoF or GoF/DN mechanisms are possible |
| 3 | *KCNQ2* | 602235 | Unlikely eligible | Missense variant in gene where LoF or GoF/DN mechanisms are possible |
| 1 | *KDM1A* | 609132 | Unlikely eligible | Missense variant in gene where LoF or GoF/DN mechanisms are possible |
| 1 | *KIF1A* | 601255 | Eligible | Missense variant in gene where both LoF and GoF/DN mechanisms are possible. Clinical evidence of successful knockdown ASO |
| 1 | *KIF5A* | 602821 | Unlikely eligible | Missense variant in gene where LoF or GoF/DN mechanisms are possible |
| 1 | *MAF* | 177075 | Unlikely eligible | Missense variant in gene where LoF or GoF/DN mechanisms are possible |
| 1 | *MAP3K7* | 602614 | Unlikely eligible | Missense variant in gene where LoF or GoF/DN mechanisms are possible |
| 1 | *MAPK8IP3* | 605431 | Unlikely eligible | Missense variant in gene where LoF or GoF/DN mechanisms are possible |
| 1 | *MAST1* | 612256 | Likely eligible | In-frame deletion in gene where GoF or DN mechanism are cause of disease, but not functionally proven for specific variant(s) in our cohort |
| 1 | *MTOR* | 601231 | Likely eligible | Missense variant in gene where GoF or DN mechanism are cause of disease, but not functionally proven for specific variant(s) in our cohort |
| 2 | *NR2F1* | 132890 | Unlikely eligible | Missense variant in gene where LoF or GoF/DN mechanisms are possible |

| | | | | |
|---|---|---|---|---|
| 1 | *NR2F2* | 107773 | Unlikely eligible | Missense variant in gene where LoF or GoF/DN mechanisms are possible |
| 1 | *POLR2A* | 180660 | Unlikely eligible | Missense variant in gene where LoF or GoF/DN mechanisms are possible |
| 1 | *POLR3B* | 614366 | Likely eligible | Missense variant in gene where GoF or DN mechanism are cause of disease, but not functionally proven for specific variant(s) in our cohort |
| 1 | *PPM1D* | 605100 | Likely eligible | Missense variant in gene where GoF or DN mechanism are cause of disease, but not functionally proven for specific variant(s) in our cohort |
| 1 | *PPP2R5D* | 601646 | Likely eligible | Missense variant in gene where GoF or DN mechanism are cause of disease, but not functionally proven for specific variant(s) in our cohort |
| 1 | *PPP3CA* | 114105 | Unlikely eligible | Missense variant in gene where LoF or GoF/DN mechanisms are possible |
| 1 | *PRKAR1B* | 176911 | Unlikely eligible | Missense variant in gene where LoF or GoF/DN mechanisms are possible |
| *5* | *PTPN11* | 176876 | Unlikely eligible | Missense variant in gene where LoF or GoF/DN mechanisms are possible |
| 1 | *RERE* | 605226 | Unlikely eligible | Missense variant in gene where LoF or GoF/DN mechanisms are possible |
| 1 | *RNF213* | 613768 | Likely eligible | Missense variant in gene where GoF or DN mechanism are cause of disease, but not functionally proven for specific variant(s) in our cohort |
| 2 | *RYR1* | 180901 | Likely eligible | Missense variant in gene where GoF or DN mechanism are cause of disease, but not functionally proven for specific variant(s) in our cohort |
| 1 | *SATB2* | 608148 | Unlikely eligible | Missense variant in gene where LoF or GoF/DN mechanisms are possible |
| 3 | *SCN2A* | 182390 | Eligible | Missense variant in gene where both LoF and GoF/DN mechanisms are possible. Clinical evidence of successful knockdown ASO |
| 1 | *SCN8A* | 600702 | Eligible | Missense variant in gene where both LoF and GoF/DN mechanisms are possible. Pre-clinical evidence of successful knockdown ASO |
| 2 | *SMARCA2* | 600014 | Likely eligible | Missense variant in gene where GoF or DN mechanism are cause of disease, but not functionally proven for specific variant(s) in our cohort |
| 1 | *SMARCA4* | 603254 | Unlikely eligible | Missense variant in gene where LoF or GoF/DN mechanisms are possible |
| 1 | *SMC1A* | 300040 | Unlikely eligible | Missense variant in gene where LoF or GoF/DN mechanisms are possible |
| 1 | *SPTBN2* | 604985 | Likely eligible | Inheritance associated with GoF or DN variants, though no functional evidence for variant(s) in our cohort |
| 1 | *TUBA1A* | 602529 | Unlikely eligible | In-frame deletion where both LoF and GoF/DN mechanisms are possible |
| 1 | *TUBB3* | 602661 | Likely eligible | Missense variant in gene where GoF or DN mechanism are cause of disease, but not functionally proven for specific variant(s) in our cohort |
| 1 | *ZBTB20* | 606025 | Unlikely eligible | Missense variant in gene where LoF or GoF/DN mechanisms are possible |

DN, dominant negative; GoF, gain of function; LoF, loss of function; OMIM, Online Mendelian Inheritance in Man

**Supplemental Table 8. Disease/condition- and delivery-related considerations for eligible and likely eligible variants identified in this study.**

| Gene symbol | Relevant OMIM phenotype (Phenotype #) | Selected condition-related considerations[a] | Possible therapeutic goals | Well-established ASO delivery to target organ(s)[e] |
|---|---|---|---|---|
| Grey rows indicate variants that were considered unlikely candidates for bespoke ASO development based on disease/condition and/or delivery-related considerations. | | | | |
| *ABCA3* | Surfactant metabolism dysfunction, pulmonary, 3 (610921) | *Rapidly progressive congenital lung disease* | Delay disease progression (e.g., as a bridge to lung transplantation) | No (lung) |
| *ABCC8* | Hyperinsulinemia hypoglycemia, familial, 1 (256450) | *Congenital onset hyperinsulinism* | Decrease islet cell hyperplasia | No (pancreas) |
| *ARID2* | Coffin-Siris syndrome 6 (617808) | **DD/ID expected**<br>**Seizures in a minority**<br>*Potential morbidity from extra-CNS congenital anomalies*<br>*Psychomotor regression not expected* | Maximize development, cognition, and functioning<br>Improve seizure control | Yes (CNS) |
| *BRAF* | Cardiofaciocutaneous syndrome (115150) | **DD/ID expected**<br>**Seizures in a minority**<br>**Risk for cardiomyopathy**<br>*Potential morbidity from extra-CNS congenital anomalies*<br>*Psychomotor regression not expected* | Maximize development, cognition, and functioning<br>Improve seizure control<br>Treat cardiomyopathy | Yes (CNS); No (cardiac muscle) |
| *CBL* | Noonan syndrome-like disorder with or without juvenile myelomonocytic leukemia (613563) | **DD reported**<br>**Increased susceptibility to haematologic cancer**<br>*Potential morbidity from extra-CNS congenital anomalies*<br>*DD can be mild*<br>*Psychomotor regression not expected* | Maximize development, cognition, and functioning<br>Decrease cancer risk and/or adjuvant cancer treatment | No (bone marrow); Yes (CNS) |
| *DES* | Cardiomyopathy, dilated, 1I (604765) | **Progressive severe cardiac phenotype (arrythmia; cardiomyopathy)** | Decrease risk of cardiac complications | No (heart) |
| *DNM1L* | Encephalopathy, lethal, due to defective mitochondrial peroxisomal fission 1 (614388) | **DD/ID expected**<br>**Psychomotor regression expected**<br>**Epilepsy common**<br>**Risk for movement disorder** | Delay neurological deterioration<br>Maximize development, cognition, and functioning<br>Improve seizure control and dystonia | Yes (CNS) |
| *FGFR3* | Achondroplasia (100800)[b] | *Prenatal-onset skeletal dysplasia* | Decrease risk of condition-related complications that | No (bone) |

| | | | | |
|---|---|---|---|---|
| | | | impact quality of life or mortality (e.g., craniocervical junction, obstructive sleep apnea, middle ear dysfunction, spinal stenosis) | |
| *FGFR3* | Crouzon syndrome with acanthosis nigricans (612247) | *Congenital craniofacial malformations* | Decrease risk of condition-related complications that impact quality of life or mortality (e.g., hydrocephalus) | No (bone) |
| *FLNA* | Heterotopia, periventricular, 1 (300049)[c] | **Epilepsy**<br>*Congenital CNS anomalies (neuronal migration disorder)*<br>*Potential morbidity from extra-CNS issues (gastrointestinal, cardiovascular, respiratory, hematologic)*<br>*Psychomotor regression not expected* | Improve seizure control | Yes (CNS) |
| *GH1* | Growth hormone deficiency, isolated, type II (173100) | **Height deficit and decreased growth velocity**<br>*Good response to recombinant human growth hormone (rhGH)*<br>*Possible structural CNS anomaly (pituitary hypoplasia)* | Increase growth velocity | Yes (liver); No (bone; muscle; other) |
| *HBB* | Thalassemia, beta (613985) | **Severe anemia** | Increase normal red blood cell production | No (bone marrow) |
| *KCNH1* | KCNH1-related disorder (-)[d] | **DD/ID expected**<br>**Epilepsy common**<br>**Reports of movement disorder**<br>*Psychomotor regression not expected* | Maximize development, cognition, and functioning<br>Improve seizure control<br>Treat movement disorder | Yes (CNS) |
| *MORC2* | Developmental delay, impaired growth, dysmorphic facies, and axonal neuropathy (619090) | **DD/ID expected**<br>**Progressive findings on brain MRI**<br>**Progressive symptoms from peripheral neuropathy** | Maximize development, cognition, and functioning<br>Delay neurological deterioration | Yes (CNS); No (PNS) |
| *MYH7* | Cardiomyopathy, familial hypertrophic, 1 (192600) | *Progressive cardiomyopathy* | Delay progression of cardiomyopathy | No (cardiac muscle) |
| *PACS1* | Schuurs-Hoeijmakers syndrome (615009) | **DD/ID expected**<br>**Epilepsy common**<br>**Reports of behavioral issues (e.g., aggression)**<br>*Potential morbidity from extra-CNS (heart) congenital anomalies* | Maximize development, cognition, and functioning<br>Improve seizure control | Yes (CNS) |

| | | | | |
|---|---|---|---|---|
| | | *Potential congenital CNS anomalies*<br>*Psychomotor regression not expected* | | |
| *PCNT* | Microcephalic osteodysplastic primordial dwarfism, type II (210720) | *Prenatal-onset skeletal dysplasia*<br>*Global vascular disease* | Promote bone growth<br>Decrease risk of neuro, cardio, renal, or other vascular complications | No (bone; vasculature) |
| *PPP2R1A* | Houge-Janssens syndrome 2 (616362) | **DD/ID expected**<br>**Epilepsy common**<br>**Behavioral issues (e.g., self-injury) common**<br>**Progressive microcephaly**<br>*Reports of ventriculomegaly*<br>*Potential congenital CNS anomalies*<br>*Psychomotor regression not expected* | Maximize development, cognition, and functioning<br>Improve seizure control | Yes (CNS) |
| *PPP2R5D* | Houge-Janssens syndrome 1 (616355) | **DD/ID expected**<br>**Seizures/epilepsy common**<br>**Reports of behavioral issues (e.g., aggression)**<br>*Psychomotor regression not expected* | Maximize development, cognition, and functioning<br>Improve seizure control | Yes (CNS) |
| *PRKACA* | Cardioacrofacial dysplasia 1 (619142) | *Congenital skeletal dysplasia phenotype*<br>*Potential morbidity from congenital cardiac anomalies*<br>*Risk of early respiratory-related death* | Increase bone growth | No (bone) |
| *RAC3* | Neurodevelopmental disorder with structural brain anomalies and dysmorphic facies (618577) | **DD/ID expected**<br>**Seizures in a minority**<br>*Congenital structural CNS anomalies (neuronal migration disorder)* | Maximize development, cognition, and functioning<br>Improve seizure control | Yes (CNS) |
| *SMPD4* | Neurodevelopmental disorder with microcephaly, arthrogryposis, and structural brain anomalies (618622) | **DD/ID expected**<br>**Seizures in a minority**<br>**Progressive microcephaly**<br>**Reports of dilated cardiomyopathy**<br>*Potential morbidity from extra-CNS congenital anomalies*<br>*Potential congenital CNS anomalies* | Maximize development, cognition, and functioning<br>Improve seizure control<br>Treat cardiomyopathy | Yes (CNS); No (cardiac muscle) |
| *TPM2* | Congenital myopathy 23 (609285) | **Progressive muscle weakness** | Delay progression of muscle weakness<br>Improve motor development and functioning | No (muscle) |
| *TUBB3* | Fibrosis of extraocular muscles, congenital, 3A (600638)[b] | *Congenital muscle fibrosis*<br>*Variable additional phenotype* | Unclear | No (ocular muscle) |

| | | | | |
|---|---|---|---|---|
| *VPS13B* | Cohen syndrome (216550) | **Progressive retinochoroidal dystrophy**<br>**Acquired microcephaly**<br>**DD/ID expected**<br>**Seizures in a minority**<br>*Potential morbidity from neutropenia and recurrent infections*<br>*Psychomotor regression not expected* | Delay progression of retinal dystrophy<br>Maximize development, cognition, and functioning<br>Improve seizure control | Yes (CNS; retina) |

CNS, central nervous system; DD, developmental delay; ID, intellectual disability; OMIM, Online Medelian Inheritance in Man; PNS, peripheral nervous system

[a]Bolded features may influence ASO amenability in a positive direction, e.g., by providing a target for treatment. Italicized features may influence ASO amenability in a negative direction, e.g., by suggesting an irreversible phenotype or added treatment risk. The listed features are not comprehensive, the category assignations are subjective, and individual patient-level factors are not considered.
[b]Individual in the study cohort had additional complex phenotype(s) that prompted genome-wide sequencing and that are unexplained by the identified variant (i.e., the condition listed here represented a partial genetic diagnosis for this individual).
[c]Individual in the study cohort had two X chromosomes.
[d]Features in the study cohort individual were most in keeping with "KCNH1-related disorder."[3]
[e]See Aartsma-Rus *et al.* (2024) for details.[2]

**Supplemental Figure 1. Clinical genome-wide sequencing tests and family designs used in the diagnosis of individuals in this study cohort.**

For the quad sequencing (affected sibling pairs and biological parents), only the proband was included in our study cohort. Graph designed using Prism.

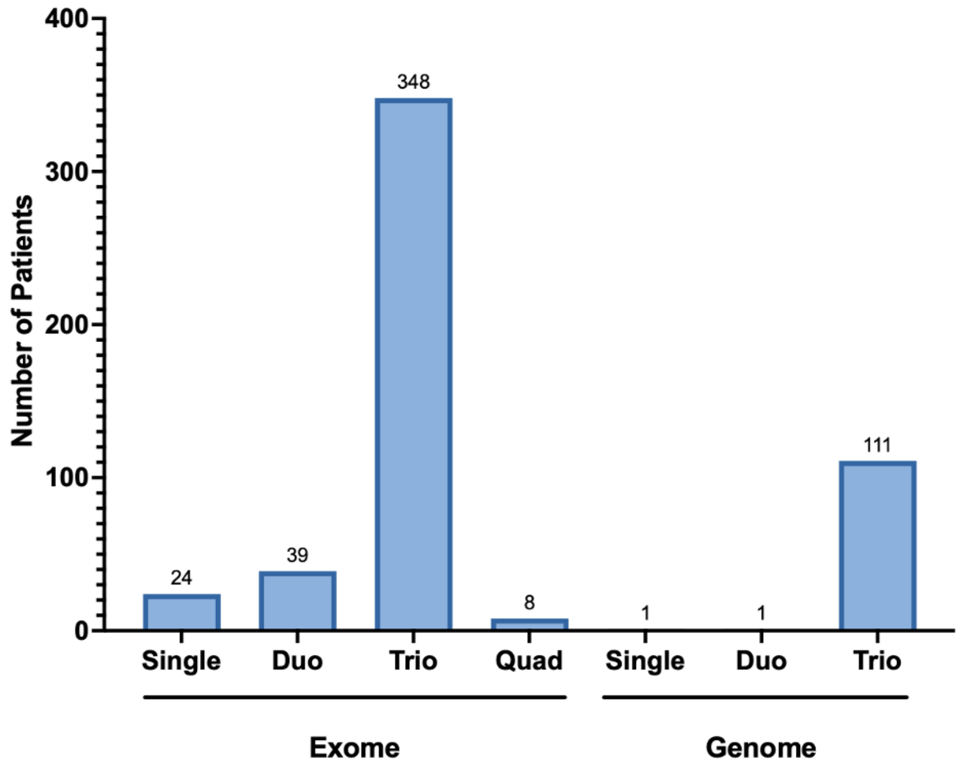

**Supplemental Figure 2. Disease category associations for genes (n=449) containing diagnostic variants in the study cohort.**

Genes were classified in "disease categories" using the Genomics England PanelApp,[4] restricted to gene-disease associations with high-level evidence. Genes can be associated with more than one disease category, and so percent total sums to greater than 100. Graph designed using Prism GraphPad.

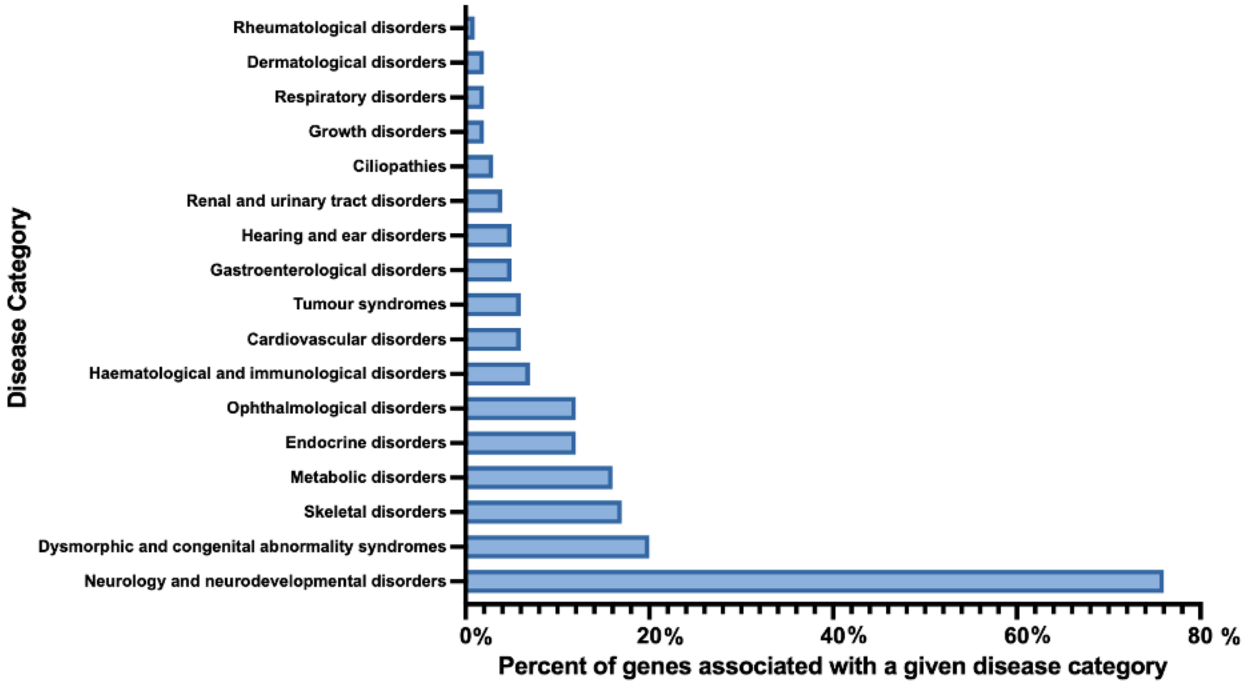

**Supplemental Figure 3. Overview of exon skipping assessment for diagnostic variants in the study cohort.**

A total of 519 variants were contained within a single exon of a gene (Supplemental Table 3). See text and N1C VARIANT guidelines[5] for details. The assessments of the remaining 18 variants are summarized in Supplemental Table 4. Additionally, 12 single or multi-exon deletions were assessed to determine whether exon skipping can restore reading frame and were classified as "not eligible" (not displayed).

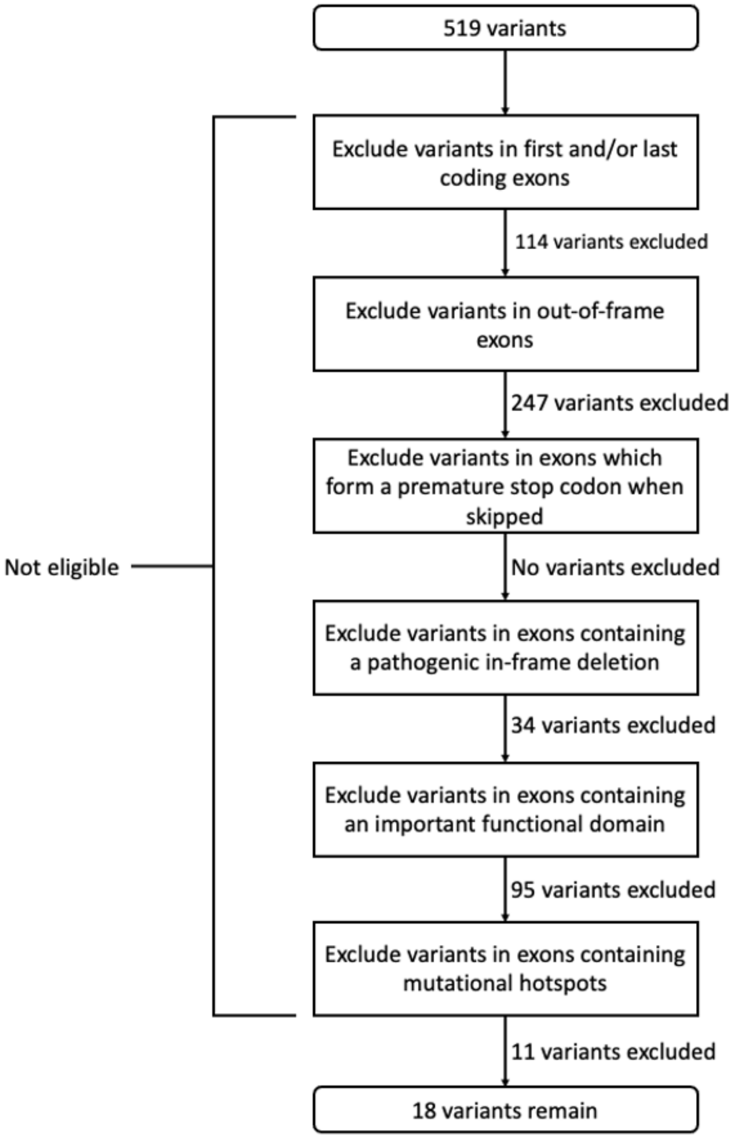

**Supplemental Figure 4. Aberrant splicing caused by *ABCA3* and *HBB* variants in the study cohort.**

The deep intronic variants NM_001089.3(*ABCA3)*:c.3863-98C>T (panel A) and NM_000518.5(HBB):c.316-106C>G (panel B) act as new donor sites, forming 150 and 165 nucleotide pseudoexons, respectively. Panel A adapted from Agrawal *et al*. 2012;[6] Panel B adapted from Treisman *et al*. 1983.[7] Exons depicted in blue, pseudoexon depicted in red, introns depicted with dotted lines. Lowercase letters represent intronic sequence, uppercase letters represent exonic sequence. Red letter indicates variant. Not drawn to scale.

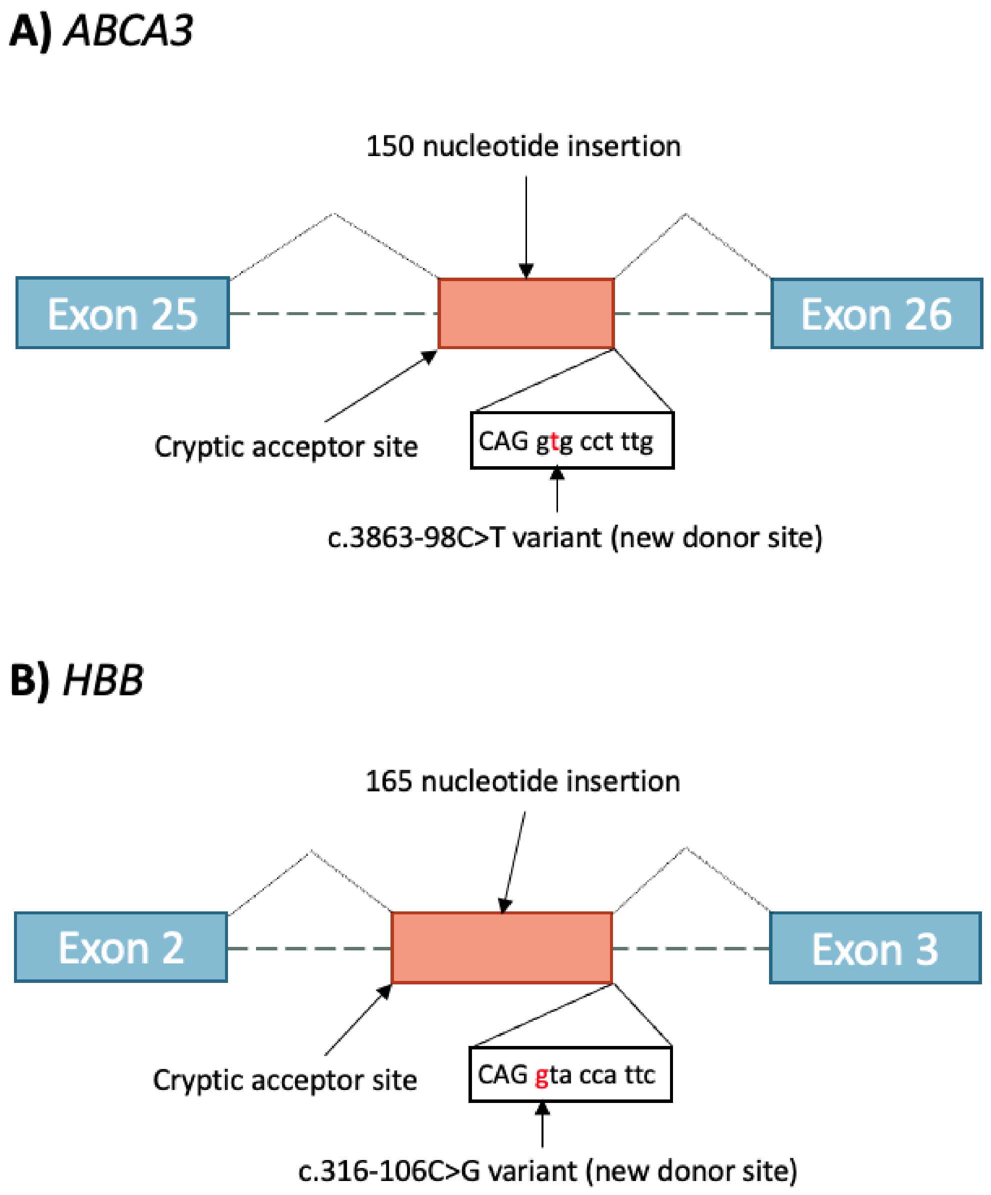

**Supplemental Figure 5. Aberrant splicing caused by an *FLNA* variant in the study cohort.**

The NM_001110556.2(*FLNA*):c.1923C>T variant acts as a new donor site, causing a 101 nucleotide deletion at the 3' end of exon 13. Figure adapted from Hehr *et al*. 2006.[8] Exons depicted in blue, deleted portion indicated in light blue, introns depicted with dotted lines. Lowercase letters represent spliced-out sequence, uppercase letters represent exonic sequence. Red letter indicates variant. Not drawn to scale.

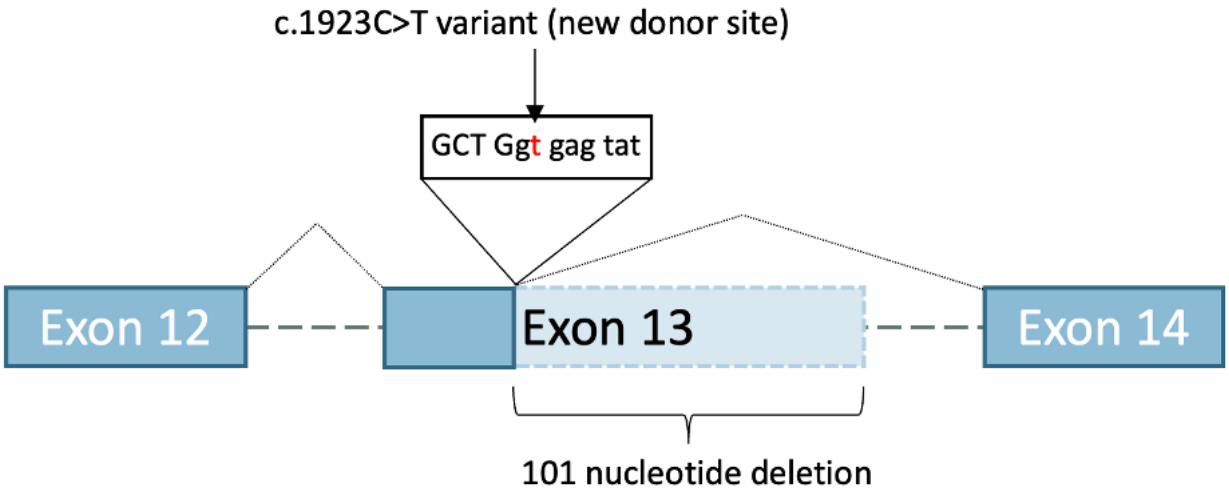

**Supplemental Figure 6. Number of variants in the study cohort for which the corresponding genes may have poison exons, naturally occurring antisense transcripts, and/or upstream open reading frames.**

Heterozygous loss-of-function variants *in trans* to a wildtype allele were assessed for whether poison exons (PEs), naturally occurring antisense transcripts (NATs), and upstream open reading frames (uORFs) are in the HUGO database (https://www.genenames.org/)[9] or in any of the four papers[10-13] discussed in the N1C VARIANT Guidelines.[5] Graph created using Prism GraphPad. Assessment of variants for eligibility towards wild-type upregulation is limited in version 1.0 of the N1C VARIANT guidelines.[5] Experimental validation and proper characterization of the role of the PEs, NATs, and uORFs is required. Not all of these elements act in an inhibitory manner to downregulate wildtype alleles, and therefore must be followed up with functional studies. One would also need to assess whether these elements are expressed at high-enough levels in the target tissue of interest to recapitulate the wildtype phenotype.

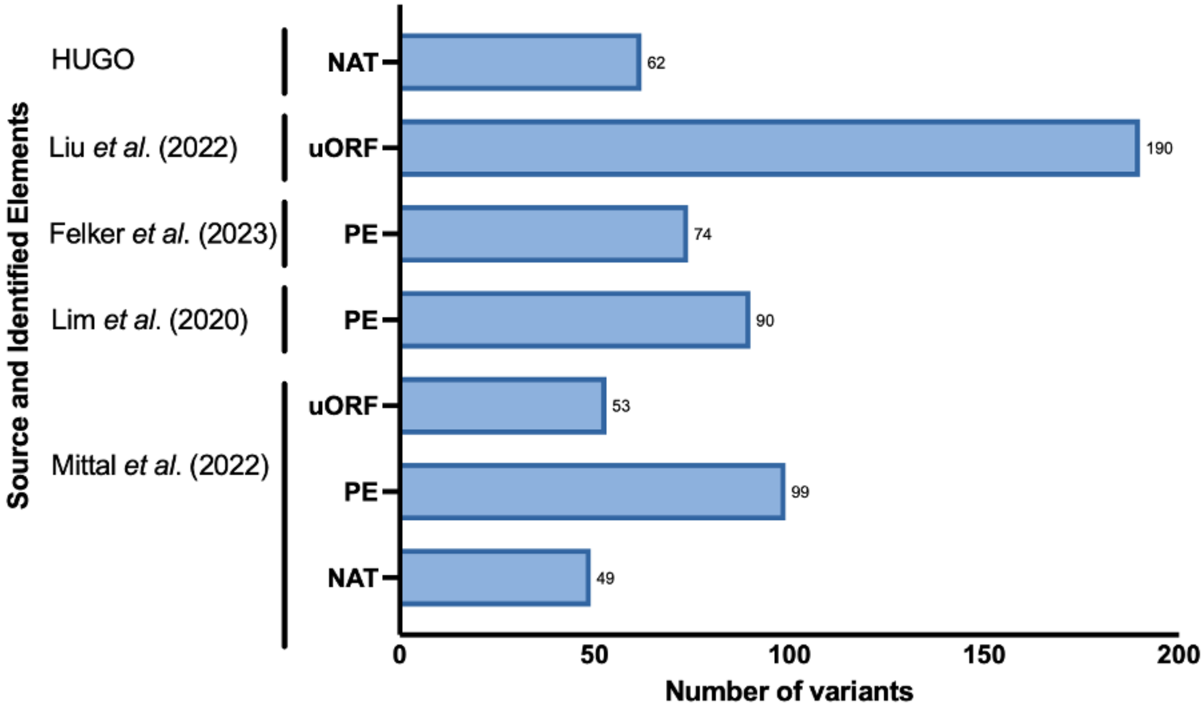

### SUPPLEMENTAL REFERENCES

1. Stern S, Wange RL, Rogers H. An Evaluation of First-in-Human Studies for RNA Oligonucleotides. Nucleic Acid Ther. 2024;34(6):276-284.
2. Aartsma-Rus A, Collin RWJ, Elgersma Y, Lauffer MC, van Roon-Mom W. Joining forces to develop individualized antisense oligonucleotides for patients with brain or eye diseases: the example of the Dutch Center for RNA Therapeutics. Ther Adv Rare Dis. 2024;5:26330040241273465.
3. Fukai R, Saitsu H, Tsurusaki Y, et al. De novo KCNH1 mutations in four patients with syndromic developmental delay, hypotonia and seizures. J Hum Genet. 2016;61(5):381-387.
4. Martin AR, Williams E, Foulger RE, et al. PanelApp crowdsources expert knowledge to establish consensus diagnostic gene panels. Nat Genet. 2019;51(11):1560-1565.
5. Cheerie D, Meserve M, Beijer D, et al. Consensus guidelines for eligibility assessment of pathogenic variants to antisense oligonucleotide treatments. medRxiv. 2024:2024.2009.2027.24314122.
6. Agrawal A, Hamvas A, Cole FS, et al. An intronic ABCA3 mutation that is responsible for respiratory disease. Pediatr Res. 2012;71(6):633-637.
7. Treisman R, Orkin SH, Maniatis T. Specific transcription and RNA splicing defects in five cloned beta-thalassaemia genes. Nature. 1983;302(5909):591-596.
8. Hehr U, Hehr A, Uyanik G, Phelan E, Winkler J, Reardon W. A filamin A splice mutation resulting in a syndrome of facial dysmorphism, periventricular nodular heterotopia, and severe constipation reminiscent of cerebro-fronto-facial syndrome. J Med Genet. 2006;43(6):541-544.
9. Seal RL, Braschi B, Gray K, et al. Genenames.org: the HGNC resources in 2023. Nucleic Acids Res. 2023;51(D1):D1003-D1009.
10. Lim KH, Han Z, Jeon HY, et al. Antisense oligonucleotide modulation of non-productive alternative splicing upregulates gene expression. Nat Commun. 2020;11(1):3501.
11. Mittal S, Tang I, Gleeson JG. Evaluating human mutation databases for "treatability" using patient-customized therapy. Med. 2022;3(11):740-759.
12. Felker SA, Lawlor JMJ, Hiatt SM, et al. Poison exon annotations improve the yield of clinically relevant variants in genomic diagnostic testing. Genet Med. 2023;25(8):100884.
13. Liu Q, Peng X, Shen M, et al. Ribo-uORF: a comprehensive data resource of upstream open reading frames (uORFs) based on ribosome profiling. Nucleic Acids Res. 2023;51(D1):D248-D261.
